# Hemochromatosis mutations, dementia and brain iron deposition: a prospective cohort study

**DOI:** 10.1101/2020.06.18.20134536

**Authors:** David Melzer, Janice L Atkins, Luke C Pilling, Christine J Heales, Sharon Savage, Chia-Ling Kuo, George A Kuchel, David C Steffens

**Author notes:** **Corresponding author:** Professor David Melzer, Epidemiology and Public Health Group, University of Exeter Medical School, College House, St. Luke’s Campus, Exeter, EX1 2LU, UK.

## Abstract

**Importance:** Brain iron deposition is common in dementia, but its causal significance is uncertain. The *HFE* p.C282Y homozygous mutation in European ancestry populations can lead to iron overload and hemochromatosis, mainly in males. Data on brain outcomes in homozygotes are scarce.

**Objective:** To estimate *HFE* variant associations with MRI features plus incident dementia diagnoses during follow-up in a large community based cohort.

**Design:** UK Biobank cohort with follow-up in routine hospitalization records (mean 8.8 years). MRI imaging available on a participant subset scanned 2014 to 2018.

**Setting:** Community cohort participants across England, Wales and Scotland.

**Participants:** European ancestry participants (n=451,186) aged 40 to 70 years at baseline, including 2,890 p.C282Y homozygotes (predominantly without baseline haemochromatosis diagnoses). MRI scanning on 9,464 males and 10,475 females, including 40 male and 75 female p.C282Y homozygotes.

**Exposure:** *HFE* C282Y and H63D genetic variants

**Main outcome and measures:** Brain MRI site specific T2* measures (lower values associated with iron deposition) and gray matter volumes. Incident dementia diagnoses during follow-up.

**Results:** Male p.C282Y homozygotes had lower T2* measures in several brain areas including the thalamus (beta = -1.04 standard deviations, 95% CI -1.33 to -0.76, multiple testing adjusted p-value=4.9*10^-10^), putamen and hippocampus, compared to those without *HFE* mutations. Male homozygotes also had smaller gray matter volumes in the putamen (beta -0.80 sd, 95%CI -1.12 to - 0.47, adjusted p=2.2*10^-4^) and ventral striatum.

Diagnoses of incident dementia (Hazard Ratio HR=2.27; 95% CI 1.36 to 3.80, p=0.002) were more common in p.C282Y homozygous men, as were delirium diagnoses (HR=2.04, CI 1.09 to 3.82, p=0.03), but there was no association with Stroke.

In p.C282Y homozygote females and p.C282Y/H63D heterozygotes, MRI associations were less marked.

**Conclusion and Relevance:** In a community sample, men with the *HFE* p.C282Y homozygote genotype had more brain iron deposition, smaller specific gray matter volumes, and increased incidence of dementia. As iron overload in hemochromatosis is treatable, early intervention may prevent or limit related brain pathology in male *HFE* p.C282Y homozygotes.

**Key Points:** *Question:* Is the hemochromatosis *HFE* p.C282Y homozygous variant in men associated with brain MRI features and incident dementia?

*Findings:* On MRI, p.C282Y homozygote males had evidence of more iron deposition in areas including the thalamus, putamen and hippocampus, plus smaller putamen gray matter volumes, compared to men without *HFE* mutations. In 451,186 UK Biobank participants during the mean 8.8 year follow-up, incident dementia diagnoses were more than twice as common in the 1,294 homozygous men.

*Meaning:* As iron overload in hemochromatosis is treatable, early intervention may prevent or limit related brain pathology in male *HFE* p.C282Y homozygotes.

## Introduction

Iron has been linked to dementia and other neurodegenerative diseases, with roles in oxygen transport, myelin production plus synthesis and metabolism of neurotransmitters^1–3^. Several rare genetic ‘Neurodegeneration with Brain Iron Accumulation’ syndromes have been described^4^. Iron accumulation in specific brain areas is found in Alzheimer’s disease (the most common form of dementia), beyond normal accumulations seen in aging^5^, and these iron accumulations are associated with amyloid plaques and tau aggregation^5^. Chelator treatment to remove iron has shown promise in Alzheimer’s disease animal models^5^ and in human randomized trials, with some evidence of slowing of cognitive decline^6^.

The homeostatic iron regulator *‘HFE’* gene p.C282Y variant relaxes controls iron absorption from the gut, leading to moderately raised serum iron levels^7^ in heterozygotes. In homozygote males especially, the variants can result in iron overload and iron deposition in many tissues ^8^. Male p.C282Y homozygotes are at substantially increased risks of liver cirrhosis and liver carcinomas, arthritis, osteoporosis, pneumonia and diabetes^9^. Iron overload in p.C282Y homozygote women is less common ^7^, likely due to iron losses in menstruation, and iron overload is unusual in p.C282Y/H63D compound heterozygotes^9^. In northern European Ancestry populations, the p.C282Y variant is carried by 10-15%, with approximately 1 in 150 (0.67%) being homozygous^10^. In North America, p.C282Y homozygosity prevalence is 0.44% in non-Hispanic whites, but much less common in other ancestry groups^7^. Iron overload in hemochromatosis is prevented and treated with venesection^8^, but many patients are currently diagnosed only after irreversible morbidity has developed.

There have been several reports of associations between *HFE* genotypes and dementia, but a recent multi-study case-control meta-analysis of the p.C282Y mutation rs1800562 and Alzheimer’s disease reported no association. However, this analysis tested overall genotype associations (which are dominated by the large numbers of heterozygotes) and did not analyze homozygote males separately^11^. It therefore remains unclear whether p.C282Y homozygotes have increased risks for brain outcomes.

Given the paucity of evidence, we aimed to test associations between *HFE* p.C282Y and p.H63D status and; 1) brain features in the MRI subset, and 2) incident dementia recorded during hospitalization in the overall sample. We used data from UK Biobank (UKB) European descent participants (n=451,186), including 16,534 MRI imaging volunteers. We hypothesized that p.C282Y homozygous males would be at most risk from brain iron deposition plus dementia during follow-up, as this group has substantially higher risks of other hemochromatosis morbidities^9^ compared to women p.C282Y homozygotes or p.C282Y/H63D heterozygotes. UKB consent does not allow individual feedback of genotypes, so the medical care and records used in the analyses were not altered by UKB identified genotypes.

## Methods

### Ethical approval

UKB ethical approval was from the North West Multi-Centre Research Ethics Committee. The current analysis was approved under UKB application 14631 (PI David Melzer).

### Data and Participants

UKB included 502,634 volunteers aged 40 to 70 years old at recruitment, living near 22 assessment centers in England, Scotland and Wales^12^. Baseline assessments (2006 to 2010) included disease history^12^ and participants consented to genotyping, plus record linkage to the UK National Health Service hospitalization routine datasets. As *HFE* p.C282Y mutations are common only in European ancestry groups^7^, we used data for the 451,186 European ancestries participants with *HFE* C282Y (rs1800562) genotype information, and also for *HFE* p.H63D (rs1799945): see Supplementary Methods for details. Data for MRI analyses were from participants who met the above criteria and volunteered for MRI (scanned May 2014 to 2018): main analyses included 19,944 such participants, with an additional analysis of data from the 16,534 with full data on all derived brain MRI measures.

### Outcomes

Magnetic Resonance Imaging (MRI) phenotypes were from the UKB brain image-processing pipeline^13^. A standard Siemens Skyra 3T running VD13A SP4, with Siemens 32-channel RF receive head coil was used. Measures included T2* signal loss, an indicator of magnetic susceptibility influenced mainly by iron in deoxyhemoglobin, storage proteins and myelin^14^. Given strong correlations between right and left hemisphere T2* iron deposition measures (correlation 0.48 to 0.79, all p<0.0001), mean values for right and left hemisphere matching variables were used in primary analyses, to reduce multiple statistical testing.

### Diagnoses and Follow-up

Incident diagnoses were from UKB follow-up hospitalization routine datasets to March 2017 for England (Hospital Episode Statistics), October 2016 for Scotland (Scottish Morbidity Record) and February 2016 for Wales (Patient Episode Database for Wales). The maximum inpatient follow-up was 11.0 years, mean 8.0 years. Incident diagnoses were ascertained from recorded International Classification of Disease 10^th^ revision codes. We used F00*; F01*; F02*; F03*; G30* to ascertain dementia diagnoses. We also ascertained diagnoses of Mild Cognitive Impairment (MCI; ICD-10 codes: F06.7), delirium (ICD-10 codes: F05*) and stroke or Transient Ischemic Attack (TIA) (ICD-10 codes: G45-G46*; I61*; I63*), as these conditions are correlated with dementia risk. Analyses of each incident condition excluded individuals with the respective prevalent diagnoses at baseline, based on participant responses at baseline interview plus ICD-10 coded hospitalization records from 1996 to baseline interview (see Supplementary Methods). For example participants who reported having doctor diagnosed dementia at the baseline interview and those who had hospitalization data with dementia diagnostic codes before baseline interview were excluded from incident dementia analyses. Dementia diagnosis accuracy has been validated in English hospital records (sensitivity 78%, specificity 92%)^15^ and Scottish routine data (positive predictive value 87%)^16^. In our preparatory analyses, *Apolipoprotein E (ApoE)* e4/e4 type was strongly associated with the recorded incident dementia diagnoses (OR=7.5 95% CI: 6.4 to 8.7, versus e3/e3), as expected.

### Statistical analysis

Given that *HFE* p.C282Y homozygotes (mainly men) are at most risk for iron overload, followed by p.C282Y/H63D, analyses and reporting focusses on men and women with these genotypes, compared to participants with neither p.C282Y nor p.H63D mutations. See Supplementary Methods for details and Supplementary Table 1 for participant numbers for all genotype groups.

For MRI analyses, we hypothesized that T2* iron deposition associated measures were most likely to be associated with *HFE* genotypes, followed by gray matter volumes. However, to be conservative and complete, we analyzed all available UK Biobank produced MRI measures, with Benjamini-Hochberg adjustment for multiple testing applied across all imaging phenotype associations (n=481 tests in p.C282Y homozygotes), to limit the false discovery rate. Linear regression was used to test associations between *HFE* C282Y & H63D genotypes and MRI measures (after calculating mean left and right measures as above), compared with having neither mutations, in males and females separately. MRI measures were quantile normalized and z-transformed before analysis.

Cox proportional hazards regression was used to test genotype associations with incident diagnoses. All models were stratified by sex, and adjusted for population genetics sub-structure using the first ten principal components generated in European-descent participants, genotyping microarray (Affymetrix Axiom array for 90% and Affymetrix BiLEVE array for 10% of participants, sharing >95% content), assessment center, plus age at assessment. There were no violations of proportional hazards assumptions for principal outcome (dementia and Parkinson’s) in Cox models. Analyses used Stata v14.1, and ‘stcox’ function for Cox models. In sensitivity analysis, we also excluded one randomly selected participant from each pair of participants related to the third degree (n=71,638) leaving 379,548 unrelated participants for sensitivity analyses. This was to avoid possible inflation of associations from family relatedness.

Missing data: We excluded the relatively small number of participants without imputed genotypes (n=15□233/502□642, 3.0%) and those with imprecise p.C282Y imputation (n=183/487□409, 0.04%). UK Biobank derived MRI gray matter volume measures were available for slightly more participants than for T2* measures (Table 2): we therefore provide analyses of all participant data for each brain MRI separate measure, and additional analyses including only participants with a full set of all MRI measures. Diagnosis data during follow-up were from all available hospital inpatient record data.

**Table 1:** Characteristics of participants included in the imaging and disease analyses, by HFE genotype and sex

| Characteristics of participants | Male |  |  | Female |  |  |
| --- | --- | --- | --- | --- | --- | --- |
|  | Wild type | C282Y+ /H63D+ | C282Y +/+ | Wild type | C282Y+ /H63D+ | C282Y +/+ |
| **Diagnosis study** |  |  |  |  |  |  |
| Number of participants | 122,860 | 4,955 | 1,294 | 145,719 | 5,746 | 1,596 |
| Age (baseline) – mean (sd) | 57.0 (8.1) | 56.9 (8.1) | 56.8 (8.2) | 56.6 (7.9) | 56.5 (7.9) | 56.9 (8.0) |
| Hemochromatosis diagnosis (baseline) | 30 | 29 | 156 | 8 | 12 | 54 |
| **Incident diagnoses\*** |  |  |  |  |  |  |
| Dementia | 631 | 32 | 15 | 487 | 19 | 6 |
| MCI | 102 | 6 | 3 | 68 | 2 | 3 |
| Delirium | 461 | 21 | 10 | 319 | 10 | 7 |
| Stroke/TIA | 1,907 | 89 | 27 | 1,351 | 45 | 11 |
| **MRI subgroup** |  |  |  |  |  |  |
| Total number | 5,694 | 253 | 40 | 6,204 | 237 | 75 |
| Age – mean (sd) | 63.4 (7.5) | 63.3 (7.7) | 62.4 (8.6) | 62.1 (7.3) | 62.3(7.0) | 63.3(7.2) |

\*Excluding diagnoses at baseline: some participants had more than one diagnosis. See Supplementary Table 1 for other genotype groups.

**Table 2:** Brain MRI median T2* and gray matter volume associations from linear regression comparing HFE p.C282Y homozygote participants to controls with neither p.C282Y nor p.H63D, in males and females separately

| Outcome description | beta<br>(standard<br>deviations) | 95% CI<br>(lower) | 95% CI<br>(upper) | Unadjusted<br>p-value | Multiple<br>testing<br>adjusted<br>p-value |
| --- | --- | --- | --- | --- | --- |
| **Males** |  |  |  |  |  |
| *Median T2\* measures (37 homozygote vs. 5183 control)* |  |  |  |  |  |
| thalamus | -1.04 | -1.33 | -0.76 | 1.0E-12 | 4.9E-10 |
| putamen | -0.98 | -1.27 | -0.70 | 1.5E-11 | 3.7E-09 |
| caudate | -0.60 | -0.90 | -0.31 | 6.6E-05 | 6.4E-03 |
| accumbens | -0.49 | -0.75 | -0.23 | 1.9E-04 | 1.0E-02 |
| hippocampus | -0.42 | -0.67 | -0.17 | 9.6E-04 | 4.2E-02 |
| *Gray matter volume (40 homozygote vs. 5692 control)* |  |  |  |  |  |
| Putamen | -0.80 | -1.12 | -0.47 | 1.3E-06 | 2.2E-04 |
| Vermis IX Cerebellum | -0.75 | -1.12 | -0.38 | 6.6E-05 | 6.4E-03 |
| V Cerebellum | -0.72 | -1.08 | -0.35 | 1.2E-04 | 9.7E-03 |
| Ventral Striatum | -0.55 | -0.84 | -0.26 | 2.1E-04 | 1.0E-02 |
| Vermis X Cerebellum | -0.76 | -1.15 | -0.36 | 1.7E-04 | 1.0E-02 |
| **Females** |  |  |  |  |  |
| *Median T2\* measures (69 homozygote vs. 5753 control)* |  |  |  |  |  |
| putamen | -0.965 | -1.219 | -0.712 | 9.2E-14 | 4.5E-11 |
| thalamus | -0.776 | -0.997 | -0.555 | 6.2E-12 | 1.5E-09 |
| caudate | -0.683 | -0.955 | -0.412 | 8.3E-07 | 5.7E-05 |
| pallidum | -0.431 | -0.660 | -0.203 | 2.2E-04 | 8.4E-03 |
| *Gray matter volume (75 homozygote vs. 6203 control)* |  |  |  |  |  |
| Putamen | -0.815 | -1.065 | -0.565 | 1.7E-10 | 2.7E-08 |
| Vermis IX Cerebellum | -0.682 | -0.914 | -0.451 | 7.7E-09 | 9.4E-07 |
| Ventral Striatum | -0.602 | -0.813 | -0.391 | 2.4E-08 | 2.3E-06 |
| Vermis VI Cerebellum | -0.470 | -0.685 | -0.254 | 1.9E-05 | 1.1E-03 |
| Caudate | -0.487 | -0.734 | -0.240 | 1.1E-04 | 5.1E-03 |
| Vermis VIIIa Cerebellum | -0.434 | -0.668 | -0.200 | 2.7E-04 | 9.5E-03 |
| V Cerebellum | -0.418 | -0.645 | -0.190 | 3.2E-04 | 9.6E-03 |
| Brain-Stem | -0.304 | -0.481 | -0.128 | 7.3E-04 | 2.0E-02 |
| Thalamus | -0.383 | -0.621 | -0.145 | 1.6E-03 | 3.7E-02 |

**Note:** linear regression models for mean of left and right measures, adjusted for age, imaging assessment center, microarray, and genetic principal components 1-10. Beta = standard deviation difference in MRI imaging phenotype between C282Y homozygous group and the “wild type” group. CIs = confidence intervals. Adjusted P = Benjamini-Hochberg adjusted p-values for 481 tests. See Supplementary Tables 2 and 3 for full results with other genotype groups. See Supplementary Tables 4 and 5 for analysis of left and right hemispheres separately.

## Results

Analyses of incident diagnoses during follow-up included 451,186 UKB participants of European descent, aged 40 to 70 years at baseline (mean 56.8 years, SD 8.0)(Table 1). Homozygous p.C282Y status was present in 0.64% (2890/451,186) of the sample, with 14.2% (64,440) of the sample being heterozygous (including 10,701 p.C282Y/H63D compound heterozygotes) (see Supplementary Table 1 for details of all *HFE* genotypes, including heterozygotes). Only 12.1% (156/1294) of male and 3.4% (54/1596) of female p.C282Y homozygotes had been diagnosed with hemochromatosis (Table 1) at baseline. Data from the brain MRI subset were available for 9,464 males and 10,475 females meeting inclusion criteria (Table 1), including 40 male and 75 female p.C282Y homozygotes.

### MRI measure associations

Overall participation in MRI was slightly more common in males (OR=1.09 95%CI 1.06 to 1.12) and slightly less common with advancing age (in years, OR=0.973 CI 0.971 to 0.975). However, male p.C282Y homozygotes were less likely to participate in the MRI (3.1% of 1,294 homozygote males versus 4.6% of men with neither p.C282Y nor p.H63D mutations: OR=0.62 CI 0.45 to 0.85). Only three male p.C282Y homozygote MRI participants (of the 40, 7.5%) were diagnosed with hemochromatosis at baseline compared to 12.1% in the 1,294 homozygote males included in diagnosis analyses. There were no p.C282Y homozygote males with incident dementia in the MRI sub-sample. Female p.C282Y homozygote participation in the MRI subset was similar to participation with neither *HFE* mutation.

Data on T2* signal loss (lower measures associated with more iron deposition) were available for 37 male and 69 female homozygotes (Table 2). T2* measures were substantially lower in male p.C282Y homozygotes within the thalamus (beta = -1.04 standard deviations ‘sd’, 95% CI -1.33 to -0.76, Benjamini-Hochberg multiple testing adjusted p-value=4.9*10^−10^), plus the putamen, caudate and accumbens, compared to neither mutations. There were also lower T2* measures in the hippocampus in p.C282Y homozygote men (beta= -0.42 sd, 95%CI -0.67 to -0.17, adjusted p=0.042). For illustration (Figure 1), we present T2 fluid attenuated inversion recovery (FLAIR) images from a male homozygote with the nearest to group T2* putamen measure, versus a similar image from an age and sex-matched participant without *HFE* mutations. Smaller gray matter volumes were found in the putamen of male C282Y homozygotes (beta -0.80 sd, 95%CI -1.12 to -0.47, adjusted p=2.2*10^−4^), and gray matter volumes were smaller in several cerebellar regions and the ventral striatum, compared to neither variants (see Supplementary Tables 2 and 3 for full results). Results of left and right hemisphere measures analyzed separately were consistent with the findings presented above (Supplementary Tables 4 and 5). Results of a sensitivity analysis in participants with complete MRI measures (Supplementary Tables 6 and 7) were also similar to those presented above.

**Figure 1:**
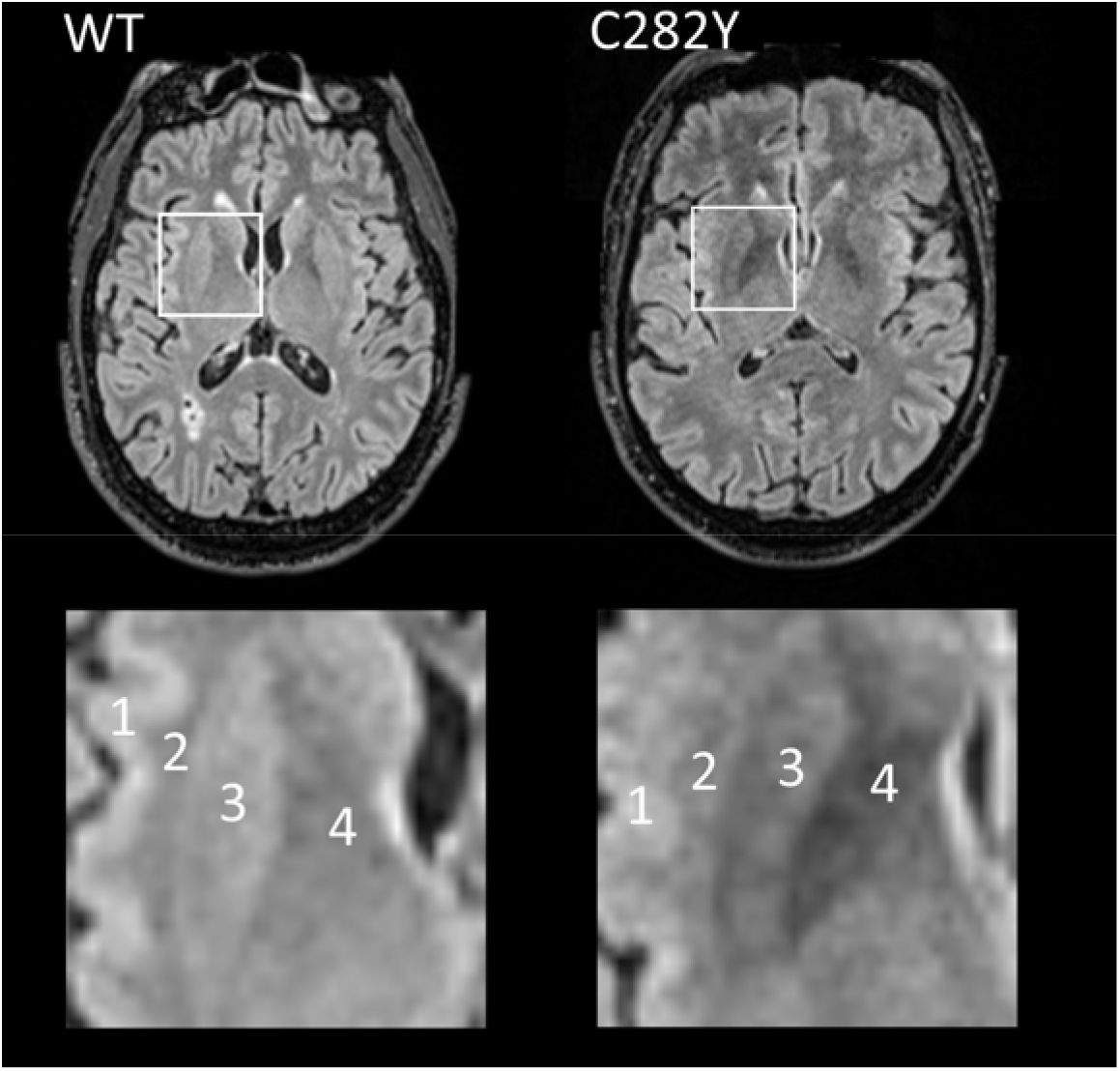
Control (left, ‘WT’) and p.C282Y homozygote (right) MRI T2 FLAIR axial images for male participants closest to respective mean putamen T2* values. In the highlighted square, cortical(1) and white matter(2) intensity appears similar in case and control but the p.C282Y homozygote image shows relative hypo-intensity (associated with iron deposition) in the putamen (3) and globus pallidus (4). Images provided by UK Biobank^©^ under license. Notes: WT = wild type or neither p.C282Y nor p.H63D mutations present. See details of image acquisition in Supplementary Information.

In women (Table 2), there were also associations between p.C282Y homozygote status and T2* measures within the putamen, thalamus, caudate, and pallidum (Table 2). Lower gray matter volumes were observed in the putamen, ventral striatum and regions of the cerebellum (adjusted p<0.05). In male and female C282Y/H63D heterozygotes and in the other *HFE* genotype groups there were similar associations, but effect sizes were substantially smaller (≤0.5 standard deviations – see Supplementary Tables 2 and 3).

### Disease associations

In the overall UKB sample there were 1,898 participants newly diagnosed with dementia during the mean 8 year follow-up (Table 1; Supplementary Table 1). Male C282Y homozygotes (0.64% of the male study population) made up 1.43% (15/1,049) of male participants with incident dementia (Table 1).

Male p.C282Y homozygotes had increased hazards for incident dementia (HR 2.27, 95% CI 1.36 to 3.80, p=0.002, compared to those with neither p.C282y nor p.H63D) (Figure 2). To remove potential bias from multiple family members, in a sensitivity analysis we included only one participant from those related to the third degree or closer (n= 174,898 after exclusions): the male homozygous association with dementia was similar (HR=2.07, 95% CI: 1.14 to 3.77, p = 0.02). To extend the evidence of association between p.C282Y homozygosity and dementia in men, we additionally tested associations with Mild Cognitive Impairment, Delirium, and Stroke or TIA diagnoses. There were increase hazards for diagnosis of incident delirium in p.C282Y homozygote males (HR 2.04, CI 1.09 to 3.82, p=0.03) and a similar but non-significant trend with Mild Cognitive Impairment (HR 2.63 CI 0.83 to 8.33). There was no association with Stroke and Transient Ischemic Attack diagnoses.

**Figure 2:**
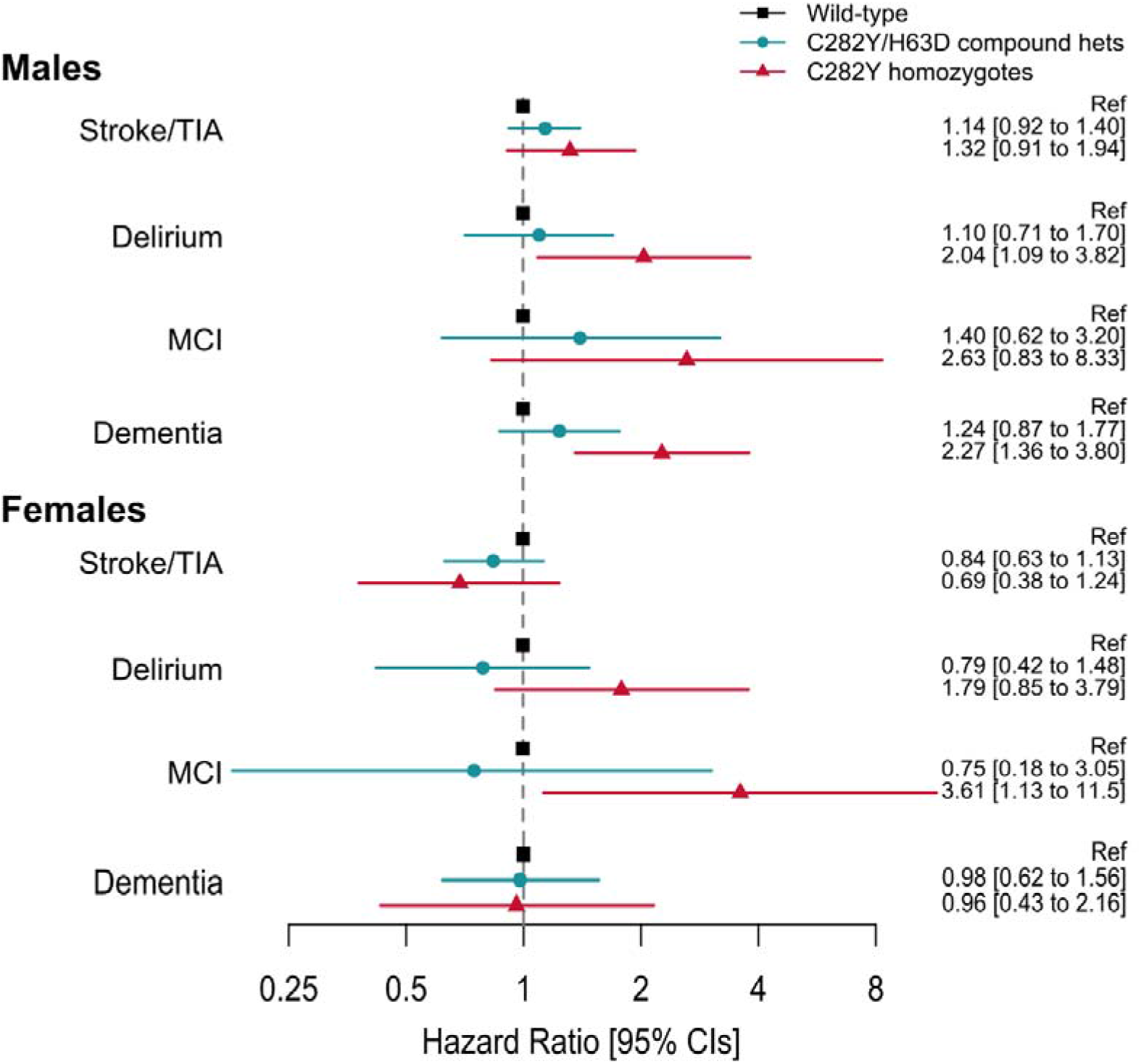
Association between main HFE genotypes and incident diagnoses: Hazard ratios (95% Confidence intervals) compared to those with no HFE mutation. **Notes:** Cox proportional hazards regression adjusted for age, imaging assessment center, microarray, and genetic principal components 1-10. Excluding each respective diagnosis at baseline. *Full results for heterozygotes are presented in Supplementary Table 8*.

In female C282Y homozygotes and the heterozygote *HFE* variants in either sex, there were no associations with dementia (Figure 2) (see Supplementary Table 8).

## Discussion

We aimed to estimate hemochromatosis genotype associations with brain MRI measures and incident dementia in the UKB community genotyped cohort. T2* signal loss occurs in the presence of magnetic susceptibility changes in the brain, caused mainly by iron^14^. In the MRI subsample, we found substantial associations in p.C282Y homozygote men with lower T2* measures (associated with greater iron deposition) in several areas involved in dementia, including the hippocampus. In addition, we found lower gray matter volumes including in the putamen and ventral striatum. In the main UKB sample (n=451,186) we found a doubling of hazard for incident dementia diagnoses during hospitalization in p.C282Y homozygote men (ascertained during the mean 8 year follow-up), plus increased hazards for delirium, for which dementia is a major risk factor^17^. Overall, these results suggest that p.C282Y homozygosity is a significant risk factor for dementia in men with European ancestries. Associations between p.C282Y heterozygous status and dementia were not statistically significant.

It is difficult to compare our findings with similar community samples with sufficient numbers of p.C282Y homozygotes because UKB contains nearly 10 times more such participants than the previously largest similar study^9^, yielding far more statistical power to detect associations. As noted, iron deposition has been linked to core Alzheimer’s disease pathologies, with iron being bound by Alzheimer’s associated amyloid β and tau, and involved in formation of oligomeric tau ^2^. A recent meta-analysis of the p.C282Y variant (HFE rs1800562, plus the transferrin variant rs1049296) found no association with Alzheimer’s disease, but no results were provided on the homozygote mutation in either men or women. In UK Biobank, a similar analysis of rs1800562 also produces no overall association (unadjusted p-value=0.021) with incident dementia, but this is because there were 64,458 heterozygotes (Supplementary table 1) and only 2890 homozygotes in analyses. An earlier meta-analysis^18^ of p.C282Y dementia association studies included data on only 18 p.C282Y homozygote cases and 37 controls and reported no association, while another meta-analysis ^19^ studied 16 p.C282Y heterozygotes and reported reduced Alzheimer’s disease risks, but included no data on homozygotes.

The brain areas with associated iron deposition in p.C282Y homozygote men included the hippocampus, a key cognitive area^20^. Iron accumulation in p.C282Y homozygote males occurred in the thalamus, an area frequently affected in early Alzheimer’s disease^21^, as well as in the putamen, where iron accumulation occurs with aging^22^ and in depression^23^. Smaller putamen gray matter volumes have been reported in neurodegenerative disorders^24^, including Alzheimer’s^20^.

## Limitations

The main limitation of this analysis is that participant ages at the end of follow-up were relatively young (mean 65.3 years old, range 40.8 to 80.9), and longer follow-up will be needed to exclude associations with dementia, especially in p.C282Y homozygous women. Dementia sub-type data are limited in UKB hospitalization records: within the male p.C282Y homozygotes with dementia, specific diagnoses were Alzheimer’s disease (n=4, ICD-10 F00.2 & ICD-10 G30.9); vascular dementia, unspecified (n=3, ICD-10 F01.9); Dementia in Parkinson’s disease (n=1, ICD-10 F02.3); dementia in other specified diseases classified elsewhere (n=1, ICD-10 F02.8) and unspecified dementia (n=6, ICD-10 F03). More work is needed to systematically assess specific dementia pathologies, including in those not hospitalized. Also, UKB volunteers tended to have somewhat less morbidity and lower prevalence of health risk factors at baseline compared to the UK population^25^, but this should have limited impact on our estimates, which are based on incident diagnoses during follow-up only in those free of dementia diagnoses at baseline. We cannot relate MRI features to incident diagnoses in the same study participants: there were no dementia diagnoses in p.C282Y homozygote men in our MRI sub-sample, likely due to the small sample size and limited follow-up time thus far, although this does have the advantage of avoiding double counting of homozygotes with dementia across the overall and MRI subset analyses.

The evidence presented of excess brain pathology is from *HFE* p.C282Y homozygote males identified through community genotyping who were predominantly not diagnosed with hemochromatosis at UK Biobank baseline. These findings add to the extensive excess morbidity reported in p.C282Y homozygote males, including excess liver disease, arthritis, diabetes, osteoporosis, and pneumonia^9^. Iron excess in p.C282Y homozygotes is usually easily and safely treated with venesection^8^. *HFE* p.C282Y associated dementia may therefore be a treatable and preventable form of dementia, but clinical trials are needed to confirm this. Our findings add to the case for systematic early ascertainment of the hemochromatosis homozygote variant, especially in men.

## Conclusions

Men with the *HFE* p.C282Y homozygous mutation developed substantially more marked brain iron deposition in dementia relevant brain areas, plus specific lower gray matter volumes. In addition, p.C282Y homozygote men were more likely to be diagnosed with dementia during the UK Biobank follow-up period. In female p.C282Y homozygotes and in heterozygotes, brain iron deposition appeared less marked, but longer follow-up will be needed to exclude associations with dementia. As the iron overload is easily treated with phlebotomy, early intervention in pC282Y homozygotes may prevent and limit the associated brain pathology and dementia diagnoses.

## Data Availability

Study data is available directly from UK Biobank following an application.

## Author contributions

DM, JLA & LCP performed the statistical analysis and drafted the manuscript. CJH advised on the MRI images. All authors were involved in design of the study, interpretation of data and revision of the manuscript.

## Acknowledgements

This research was conducted using the UK Biobank Resource, under application 14631: the authors thank the UK Biobank participants and coordinators for this unique dataset.

All authors declare no conflicts of interest.

